# Trimodality Treatment in Malignant Pleural Mesothelioma – An Ordeal or The Real Deal?

**DOI:** 10.1101/2020.05.13.20087189

**Authors:** Naveen Mummudi, Asfiya Khan, Anil Tibdewal, Rajiv Kumar, Sabita Jiwnani, George Karimundackal, CS Pramesh, Jai Prakash Agarwal

## Abstract

**BACKGROUND:** Malignant Pleural Mesothelioma is an uncommon and aggressive disease associated with asbestos exposure. Management of MPM is complex and controversial as there is paucity of good quality evidence. Multimodality treatment with surgery, systemic therapy and radiation therapy is an option in non-metastatic MPM. We intend to analyze toxicity and outcomes in patients who received trimodality treatment for non-metastatic MPM at our institution.

**METHODS and MATERIALS:** We reviewed the electronic medical records of surgically managed MPM patients at our institution in the last decade. Patient details, disease characteristics and treatment information were retrieved from the institutional electronic medical record and radiation oncology information system. Dosimetric parameters of target volume and organs at risk were documented from Eclipse workstation (v13.6, Varian medical systems). SPSS was used for statistical analysis.

**RESULTS:** Between January 2008 and October 2018, 21 patients (17 male and 4 female) underwent surgery for MPM – all but 2 patients underwent extra-pleural pneumonectomy (EPP). Primary was located in the right and left in 11 and 10 patients respectively. Epithelioid MPM was the commonest histology (17 patients - 81%). Resection was R0 in 18 patients and R2 in 2 patients. Four patients had minor complications like wound erythema, wound seroma with cellulitis and hypotension and 8 Patients had major complications like pneumonia, rib fracture, pulmonary hypertension and pulmonary stump thrombus. All patients received neoadjuvant Pemetrexed/platinum doublet chemotherapy, except for 2. Fourteen patients received adjuvant hemithoracic RT; of these, 2 underwent treatment elsewhere and 2 were treated with conventional technique. Ten patients treated with conformal technique at our institute and dosimetric data was available for analysis. Average time to start RT after surgery was 51 days (range 32-82 days). All patients were treated with conformal technique using IMRT/VMAT to a dose of 45Gy in 25 fractions; one patient received a further boost of 5.4Gy. Mean overall RT duration was 35 days (range 30 – 42 days). Acute toxicity was uncommon; Grade I/II Pneumonitis was seen in 4 patients. One patient developed grade III acute lung toxicity unrelated to RT. At a median follow up of 25 months, 8 patients developed progressive disease. Eight patients had died, of whom six died due to disease and two died in immediate post op period. Two-year DFS and OS were 58% and 73% respectively.

**CONCLUSION:** In spite of the extensive surgery and complex hemithoracic RT, we demonstrated excellent dosimetric, toxicity profile and favorable outcomes in non-metastatic MPM.

## INTRODUCTION

Malignant Pleural Mesothelioma (MPM) is an uncommon disease originating in the mesothelial surface of pleura and has strong association with asbestos exposure. Prior thoracic radiation, exposure to erionite (a mineral found in gravel road) and few genetic mutations (BRCA1-associated protein-1, BAP1 gene) have also been linked as a risk factor in the development of MPM. Mesothelioma is an aggressive malignancy as most patients present with advanced disease. Management of MPM is complicated and controversial as there is paucity of good quality evidence. Also outcomes have remained dismal despite aggressive treatment strategies with median survival of less than 2 years(1,2). Tri-modality treatment with surgery, systemic therapy and radiation therapy is an option in stage I-III MPM, but not widely adopted, in view of the complexity of the procedure, associated toxicity and absence of strong evidence to support its practice(3,4). Analysis from a hospital registry based database showed that the median survival was 4.8, 11.3, 15.3, and 19.9 months, respectively, with no therapy, chemotherapy alone, chemotherapy followed by surgery, and tri-modality therapy including radiation therapy(5).

Surgery in MPM is performed to achieve maximal cyto-reduction, as complete microscopic surgical resection is highly unlikely, and MPM almost always recurs after surgery alone. Surgical resection options in MPM includes pleural decortication, which involves removal of involved pleura along with gross tumour and extra pleural pneumonectomy (EPP) which is the en-bloc removal of involved visceral and parietal pleura, ipsilateral lung, ipsilateral diaphragm and the pericardium. Lack of robust data has made the choice between the two surgical procedures contentious(6). To improve resectability and local control rates, neoadjuvant chemotherapy is administered along with aggressive surgery and post-operative radiotherapy(4). Postoperative radiotherapy (RT) is not only an area of debate, but also extremely challenging as radiation fields have to cover a large and anatomically complex volume extending from the apex of the lung to inferiorly, the costo-diaphragmatic recesses and medially, including the diaphragmatic crus down to the lumbar vertebrae(7).The Swiss SAKK 17/04 trial is the only controlled prospective trial to investigate the role of postoperative RT as part of tri-modality therapy in MPM did not show any benefit with RT(8). However, the negative results have to be interpreted with caution because of the small number of patients, heterogeneity of RT techniques, lack of a central plan review and a high incidence of death in irradiated patients. The aggressive behaviour of the disease coupled, toxic treatment modalities and dearth of good quality evidence makes MPM one of the obstinate malignancies to treat.

At our institution, a tertiary care oncology centre, we deliver tri-modality treatment with neoadjuvant chemotherapy followed by extra-pleural pneumonectomy and adjuvant hemi-thoracic radiation in patients with non-metastatic MPM. In this report, we analyse and describe the radiation technique, dosimetric parameters, toxicity, outcome of patients who underwent trimodality treatment over the past ten years at our centre.

## MATERIALS AND METHODS

This single institutional retrospective observational study was approved by the institutional review board and informed consent was waived. From a prospectively maintained database, we reviewed patient details, disease characteristics and treatment information of surgically managed non-metastatic MPM patients treated in the last ten years at our institution. Inclusion criteria consisted of previously untreated histologically proven MPM of all subgroups, clinical stage T1-3,N0-2 disease according to AJCC 8th edition. All clinical management decisions were discussed and decided in a multidisciplinary tumour board prior to initiation of treatment.

Between January 2008 and October 2018, we identified 21 consecutive patients with non-metastatic MPM who underwent radical curative surgery. Patients underwent either an extra-pleural pneumonectomy or lung sparing surgery like pleural decortication.

Patients were planned for2-3 cycles of neoadjuvant chemotherapy with pemetrexed/platinum doublet chemotherapy. Patients received cisplatin(75mg/m2) or carboplatin (area under curve, 5) and pemetrexed 500mg/m2 every 3 weeks followed by response assessment with CECT Thorax. Following physical and pulmonary evaluation, patients were then planned for further surgical management, if fit.

### RT Technique

Patients planned for postoperative adjuvant hemi-thoracic RT were immobilized and simulated in supine position with both arms overhead. Planning CT scan was obtained from the level of cricoid cartilage till second lumbar vertebra. The entire ipsilateral hemi thorax was contoured as the clinical target volume (CTV), which included parietal and visceral pleura, diaphragmatic insertion, involved lymph node station and recesses. CTV superiorly extended from the superior border of T1 vertebral body to the insertion of diaphragm inferiorly. Laterally, the parietal pleura was covered along the ribs. Medially, the mediastinal pleural attachments were included till the hilum. PTV was generated with an isotropic 7mm margin to CTV.The dose prescribed to the Planning target volume was 45Gy in 25 fractions with boost dose to areas of high risk of failure.Dose constraints to contralateral lung were a mean lung dose less than 8Gy; V_5Gy_< 60% and V_20Gy_ of 4-10%. Treatment planning was done using AAA or a convolution/superposition algorithm. Treatment was delivered in a linear accelerator using static intensity modulated radiation therapy (IMRT) or volumetric modulated arc therapy (VMAT).

Data was retrieved from institutional electronic medical record and Radiation Oncology Information System, indigenous databases that record the course of patient history, investigations and treatment received at the institution. Radiation dosimetric parameters of target volume and organs at risk were documented from Eclipse workstation (v13.6, Varian medical systems). Toxicity was scored with the Common Terminology Criteria for Adverse Events version 4.0. Actuarial rates of disease free survival (DFS) and overall survival (OS) were estimated by using the Kaplan-Meier method and compared with log-rank tests. survival outcomes were estimated with the Kaplan-Meier method. SPSS was used for statistical analysis.

## RESULTS

Patient and demographic characteristics are listed in Table 1. All patients except for 2 received NACT; ten patients received pemetrexed and cisplatin combination while the rest received pemetrexed and carboplatin. EPP was the predominant surgery performed (90%) and only 2 underwent decortication. Complete resection was possible in 18 patients (86%). Four patients had minor post-operative complications like wound erythema, wound seroma with cellulitis and hypotension; eight patients had major complications like pneumonia, rib fracture, pulmonary hypertension and pulmonary stump thrombus. All patients had macroscopic complete resection except for 2 patients who had R+ resection. Commonest histology encountered was epithelioid mesothelioma (81%).

**Table 1.**
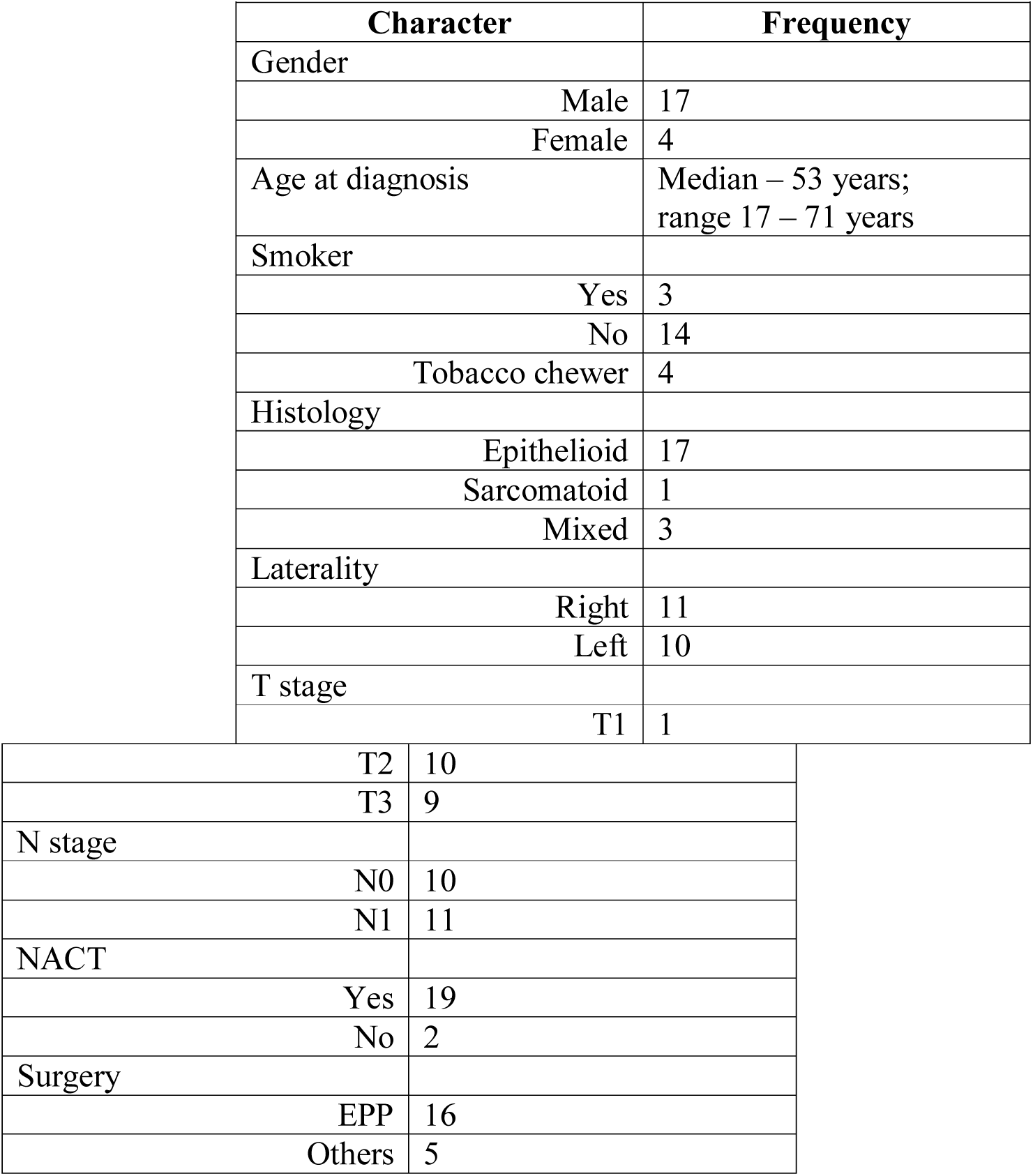
Patient characteristics (n=21)

Adjuvant hemithoracic radiation was planned in 14 patients; two patients received RT at a different centre and dosimetric parameters were not available for analysis. Two other patients received treatment using conventional antero-posterior portals and were also not included in the dosimetric analysis. All of the remaining 10 patients were treated with conformal technique; 6 patients were treated with intensity modulated radiation therapy (IMRT) and the rest treated with Volumetric modulated arc therapy (VMAT)(see Figure 1).

**Figure 1.**
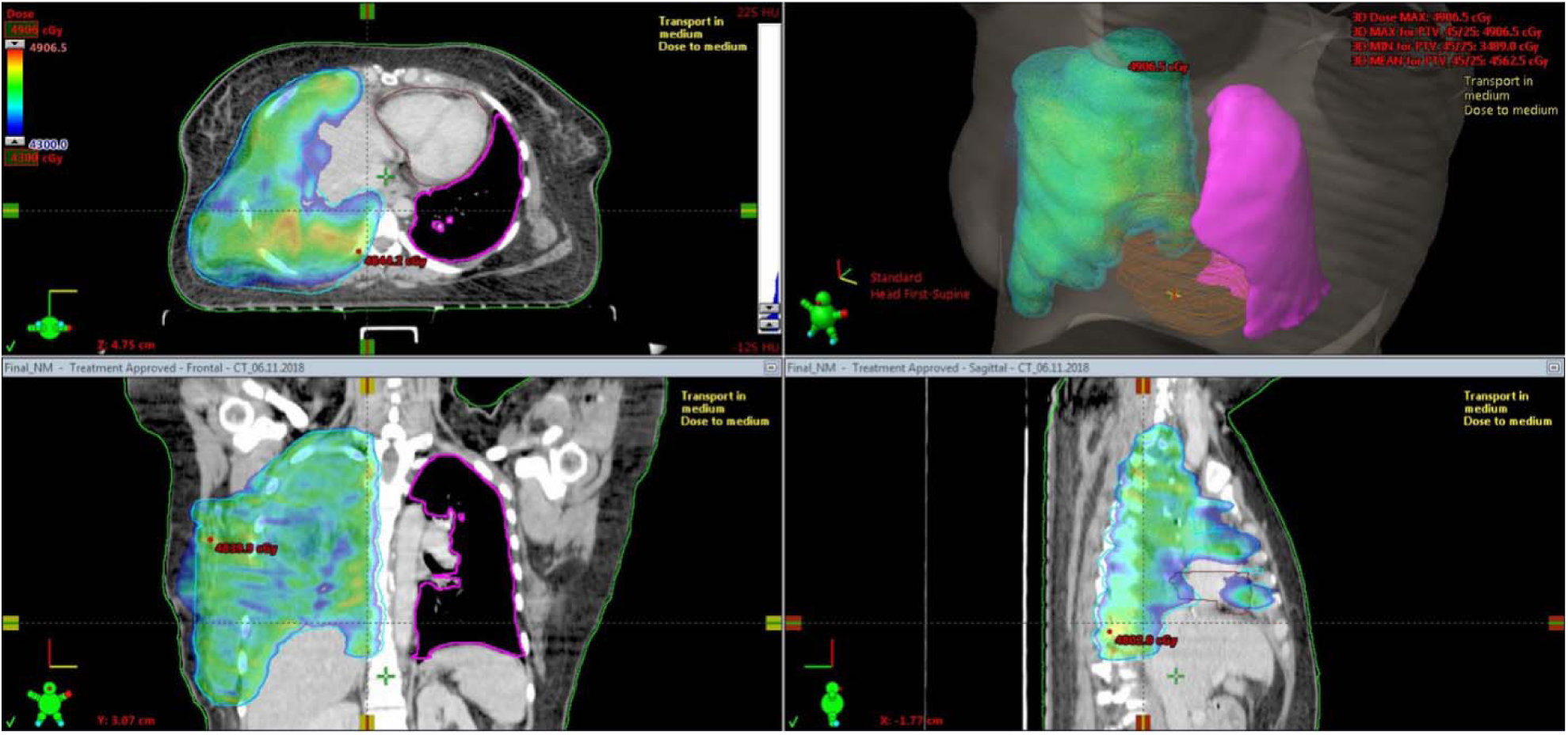
Dose distribution of a typical hemithoracic IMRT plan

Average time to start RT after surgery was 51 days (range 32-82 days). Mean dose delivered was 45Gy (range 34 to 50.4Gy) in 1.8Gy per daily fraction size. Only one patient did not complete treatment and another patient received a further 5.4Gy boost to high risk tumour bed. Mean duration of RT was 35 days (range 30-42 days).

Dosimetric characteristics are shown in Table 2. Mean Dose to contralateral Lung was 5.95Gy; volume of lung receiving 5 and 10Gy were 43% and 7.5% respectively. The mean heart dose was 20.55Gy; V_30Gy_ (%) received by heart was 26 *%*. Mean dose to liver was significantly less at 14Gy. As expected, dose received by heart was more in left sided primary whereas liver received comparatively more dose in right sided primary.

**Table 2.**
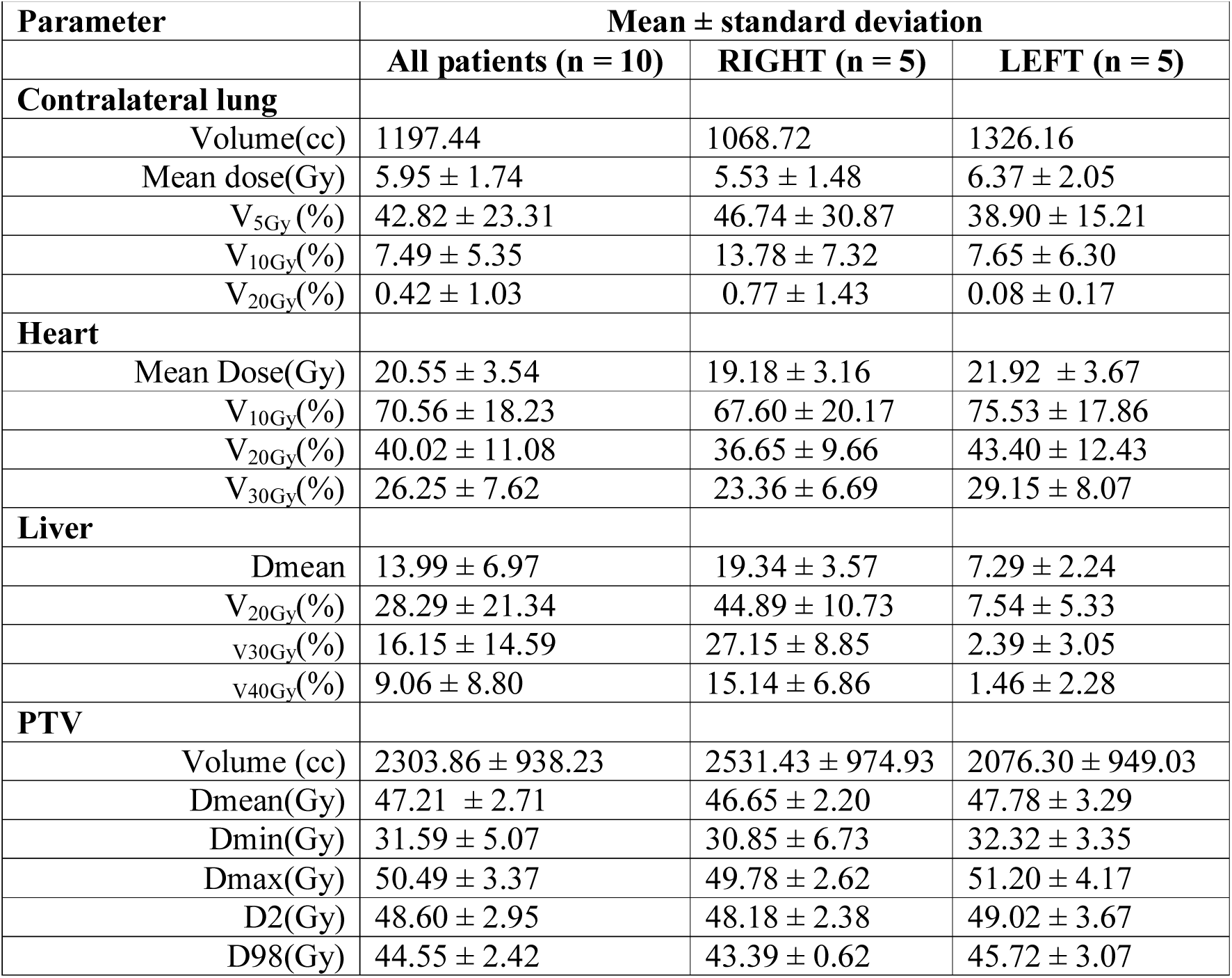
Dose parameters

### RT Toxicity (see Table 3)

Grade I oesophagitis was seen in 3 patients and one patient had Grade II oesophagitis (all left sided tumours). No patients developed grade 3/4 esophagitis. Grade I / II pneumonitis was seen in 4 patients. One patient developed Grade III acute lung toxicity and further treatment was stopped at 34Gy; however, on radiological evaluation, there was no evidence of radiation induced pneumonitis. No patient developed chronic pneumonitis or other late toxicity.

**Table 3.**
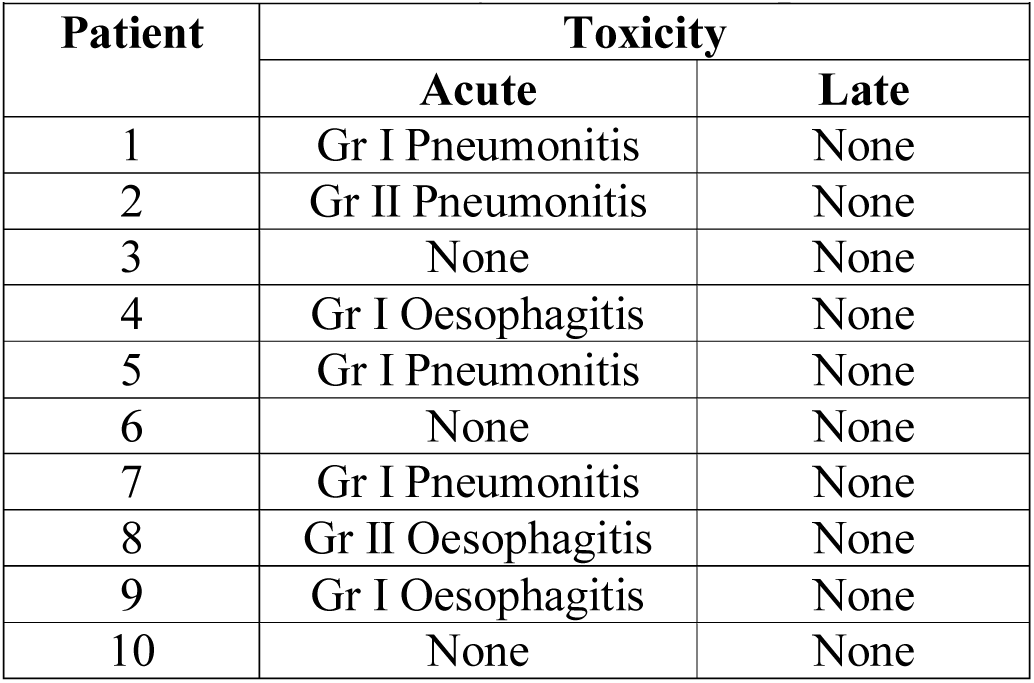
RT toxicity of individual patients

### Survival

Median follow up was 25 months (IQR 13.5-32.5 months) for the entire cohort. Eight patients developed progressive disease, of whom 2 had local disease recurrence, 3 had local and distant metastases, three more had peritoneal disease recurrence only. Of the 14 patients who received RT, there were 6 patients with disease recurrence – one with local disease only, 2 with local and distant.

At last follow up, 13 patients were alive of whom ten were alive with controlled disease; three patients had received systemic chemotherapy for metastatic disease. Eight patients had died, of whom six died due to disease and two died in the immediate post-operative period; 30-day mortality after surgery was 9.5%. In the RT cohort, there were 5 deaths, of which one alone was unrelated to the disease.

One year and 2-year disease free survival (DFS) were 65% and 58% respectively (Figure 2). Median PFS was significantly better in patients with epithelioid histology (p<0.001), but was not different for T or N stage. DFS did not differ for type of surgery, chemotherapy agents or RT delivery. Median overall survival (OS) was 41 months for the entire cohort; 1-yr and 2-yr OS were 90% and 73% respectively (Figure 3a). Patient receiving RT had a numerically better OS than those who did not receive (41 months vs 18 months; p=0.18) (Figure 3b); patients with node negative disease had a trend towards better OS (p=0.07). None of the other clinical factors was predictive for survival.

**Figure 2.**
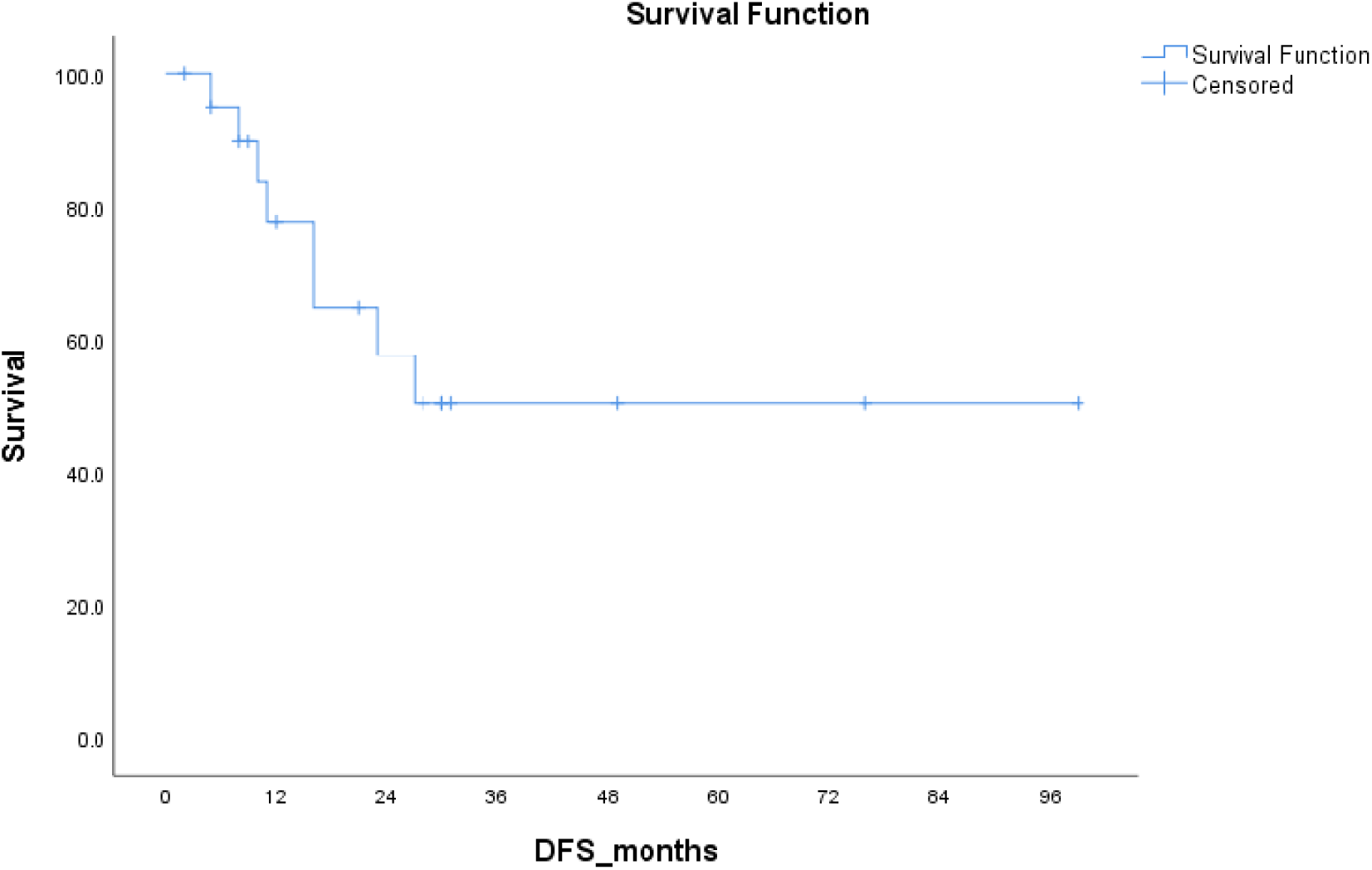
Disease free survival (DFS)

**Figure 3a.**
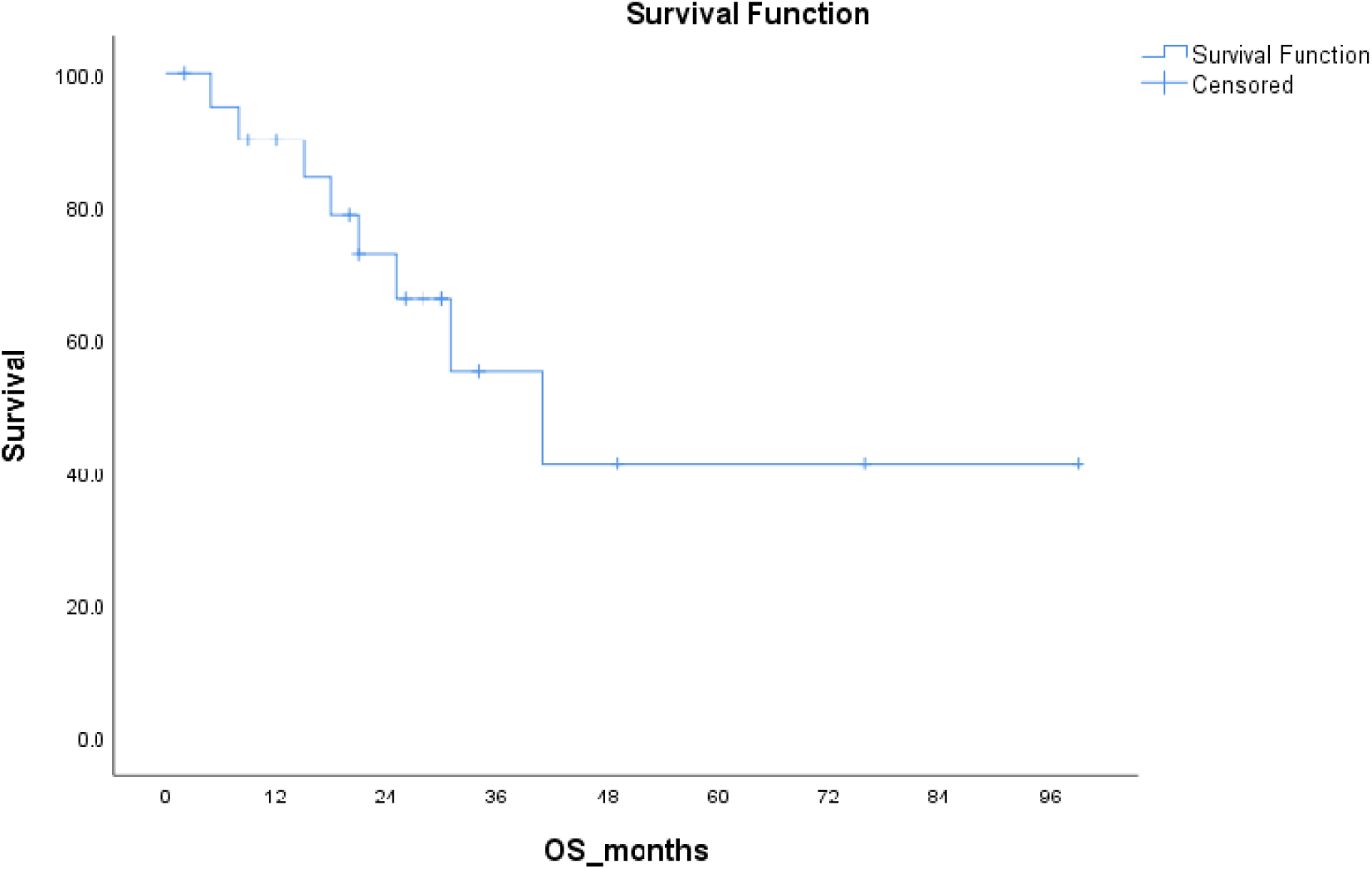
Overall survival (OS)

**Figure 3b.**
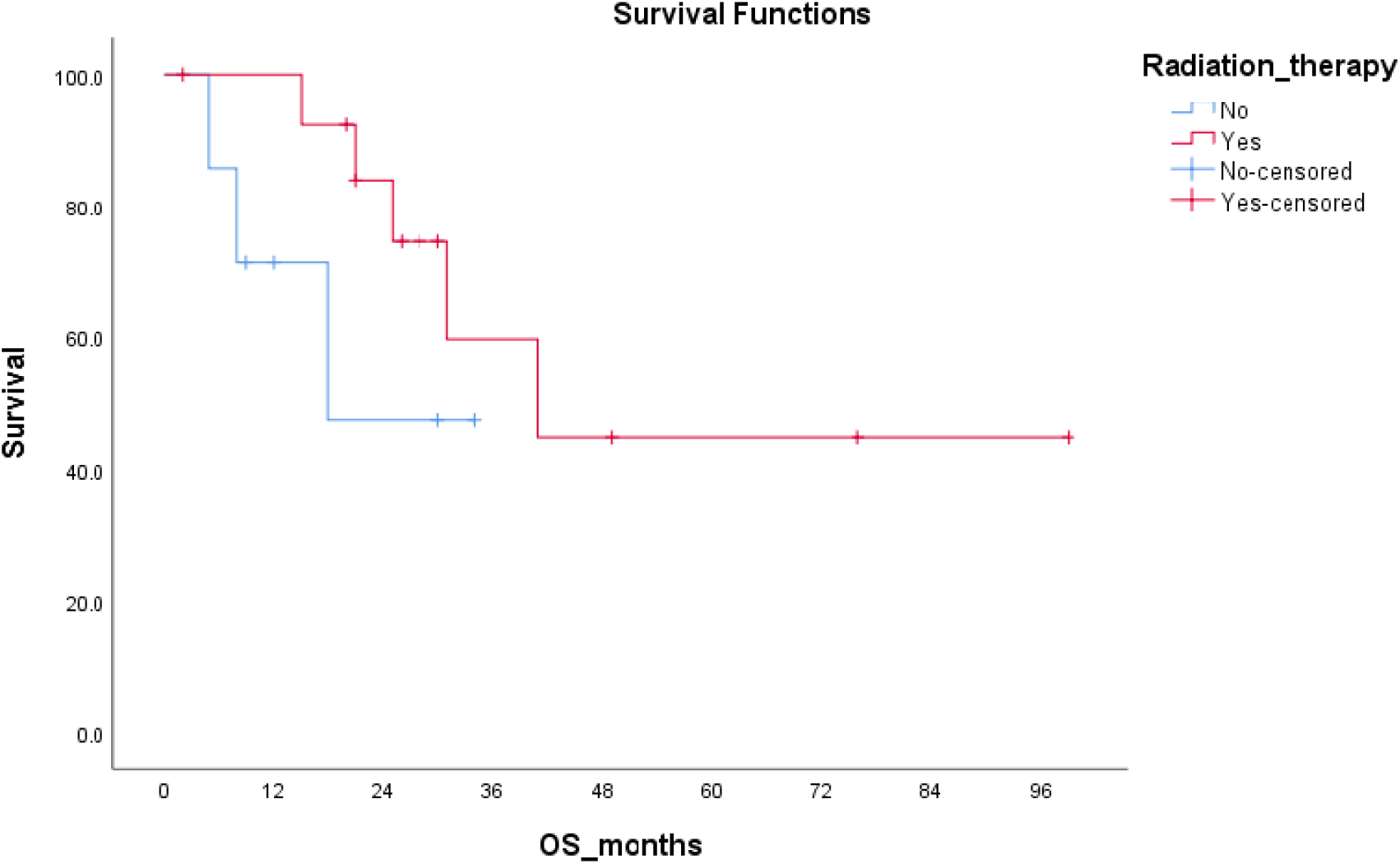
Overall survival (OS) in patients who received RT vs no RT

## DISCUSSION

Given the rarity and aggressive behaviour, the optimal management of MPM remains controversial and best managed by a multidisciplinary team with experience(9). Patients with stage I-IIIA (AJCC 8^th^ edition) are offered surgical management if they are medically fit and possess adequate pulmonary reserve. However, the choice of surgical procedure remains mired in controversy. Few meta-analysis(10-12) have shown that EPP is associated with higher morbidity and mortality compared to pleurectomy or decortication (P/D) and also have inferior survival outcome(13). MARS, a randomized study comparing EPP and P/D observed higher surgical morbidity in EPP arm. However, the study had relatively small number of patients and was intended primarily to study the feasibility of randomizing 50 patients over 1 year for surgery(6). Currently NCCN considers both surgical options as reasonable and the decision to choose one over the other dependant on factors like histology, stage, pulmonary reserve, surgical expertise, availability of adjuvant treatment options. At our institution, non-metastatic MPM patients are routinely discussed in a multidisciplinary meeting to decide on surgical and adjuvant therapy.

Adjuvant hemithoracic radiation therapy has been shown to decrease the risk of loco-regional recurrences after surgery for MPM. Data from MD Anderson Cancer Center showed that postoperative radiation therapy after EPP results in superior local control(14). In a recently published analysis of the US national cancer database (NCDB), improved survival was associated with trimodality treatment compared with surgical intervention and chemotherapy alone(15). While radiation can provide durable local control, the presence of critical structures such as the contralateral lung, heart, kidney, spleen and the complex nature of the target volume has traditionally limited the ability to safely deliver RT for patients with MPM. Presence of collection in the hemithorax, seroma in the chestwall, shift of mediastinum leads to tissue inhomogeneity, lateral electronic disequilibrium at tissue interfaces which results in significant inaccuracies in dose calculation accuracy and distribution(16).

Even with the practice of IMRT, early experience of hemithoracic RT with IMRT resulted in increased fatal pneumonitis(17), possibly because of increased “low-dose spillage” into the contralateral lung. Recent advances and technological sophistication in RT planning and delivery have resulted in increased conformity of radiation dose culminating in expanded applications of RT for patients with MPM. Compared with three dimensional conformal RT (3DCRT), IMRT has been shown to improve the planning target volume coverage(18). Although a modified electron-photon technique has the potential to minimize dose to contralateral lung and other structures when compared with IMRT(19), associated dose inhomogeneity, unpredictable target coverage may lead to increased loco-regional failure. In the ten patients we treated at our institute, we observed an average conformity index of 0.98 while the mean heterogeneity index was 0.07, highlighting excellent target coverage and dose homogeneity.

In all the cases, we were able to attain the stringent dose constraints set to the contralateral lung, liver and the kidneys. Although this could be explained by the lower total dose of 45Gy that we used, higher doses of RT is indicated only in case of macroscopic residual disease and may potentially lead to increased toxicity with no additional benefit in local control. In a study of 86 patients undergoing trimodality therapy using IMRT, the authors observed low rates of grade 3 or more esophagitis (16%) and pneumonitis (12%) without compromising the target coverage(20). We implemented strict planning guidelines with emphasis on the OAR dose constraints which has resulted in improvement in overcoming acute and late toxicities. We noted low incidence of acute and chronic clinical esophagitis and pneumonitis. The only patient in whom RT was interrupted for respiratory symptoms did not show any radiological evidence of pneumonitis at follow up. The recent patients in our cohort were treated with VMAT, which is a newer technique to deliver continuous, dynamic, modulated radiation over a rotational arc of gantry which allows for even better dose conformality than static IMRT with shorter treatment times(21,22).

Since routine use of prophylactic radiotherapy to prevent tract metastases was not shown to be beneficial post large-bore thoracic interventions in patients with metastatic MPM(23), we do not include the surgical scar in our target volume. In our series, of the three patients who failed locally after receiving RT, there was no incidence of tract site deposit.

In a multi-centre phase II randomized trial, addition of hemithoracic RT did not result in any significant difference in local-regional progression-free or overall survival (8,24). However the study was severely underpowered to detect any difference between the groups. Incidentally, we did not find any improvement in survival parameters with the delivery of RT. The two year DFS and OS for the entire cohort in our series was 58% and 73% respectively. For the patients who received RT, the corresponding figures were 61 and 84% respectively. Although statistically not significant, patients who received RT showed better outcome, with a median overall survival of 41 months. Similar to other studies, we noted better outcome in patients with epithelioid morphology(14,20).

We acknowledge the inherent weaknesses and limitations of our retrospective study. The comparison of OS and DFS outcomes in patients who received RT and those who did not is subject to bias because of underlying factors that portend a worse prognosis. The small number of patients studied over a decade long period serves only to highlight the rarity of the disease. While, adjuvant RT after EPP allows optimal dose to be delivered to the tumour bed with a lower risk of toxicity, as the ipsilateral lung is removed, recently some centres have also explored the use of lung sparing hemithoracic RT to the pleural surface alone after P/D. Studies evaluating this technique, known as intensity modulated pleural radiation therapy (IMPRINT) have reported it to be safe with no incidence of grade 3/4 pneumonitis(25). Neoadjuvant accelerated RT before surgery is also a promising approach which few centres have studied with success(26,27). Immunotherapy has already shown some potential in the systemic treatment of MPM(28-30) and hopefully would bring about a paradigm shift as it has in the management of non-small cell lung cancer.

With a lack of acceptability and consensus on optimal treatment, complex, arduous surgery and precise radiation procedures, MPM continues to baffle thoracic oncologists with sobering outcomes. Tri-modality treatment of MPM is an aggressive treatment strategy; despite the extensiveness of surgery and complexity of hemi-thoracic RT, we demonstrated excellent dosimetry, toxicity profile, favourable outcomes and promising median overall survival in non-metastatic MPM.

Although the exact and definite role of radiotherapy in the treatment paradigm of mesothelioma remains to be defined, increasing evidence supports the fact that mesothelioma is sensitive to radiotherapy treatment(22). With this account of institutional treatment outcomes, our study adds to the existing literature supporting the efficacy of trimodality therapy in the treatment of MPM, bringing about renewed hope for this rare malignancy.

## Data Availability

All data available for review.

